# Automated Prediction of Ischemic Brain Tissue Fate from Multi-Phase CT-Angiography in Patients with Acute Ischemic Stroke Using Machine Learning

**DOI:** 10.1101/2020.05.14.20101014

**Authors:** Wu Qiu, Hulin Kuang, Johanna M. Ospel, Michael D Hill, Andrew M Demchuk, Mayank Goyal, Bijoy K Menon

## Abstract

**Background and objective:** Multiphase CT-Angiography (mCTA) provides time variant images of the pial vasculature supplying brain in patients with acute ischemic stroke (AIS). Here, we develop and test a machine learning (ML) technique that predicts infarct, penumbra and tissue perfusion from mCTA source images.

**Methods:** 284 patients with AIS were included from the Prove-IT study. All patients had non-contrast CT, mCTA and CTP imaging at baseline and follow up MRI/NCCT imaging. Of the 284 patient images, 140 patient images were randomly selected to train and validate three ML models to predict infarct, penumbra, and perfusion parameter on CTP, respectively. The remaining unseen 144 patient images independent of the derivation cohort were used to test the derived ML models. The predicted infarct, penumbra, and perfusion volume from ML models was spatially and volumetrically compared to manually contoured follow up infarct and time-dependent Tmax thresholded volume (CTP volume), using Bland-Altman plots, concordance correlation coefficient (CCC), intra-class correlation coefficient (ICC), and Dice similarity coefficient (DSC).

**Results:** Within the test cohort, Bland–Altman plots showed that the mean difference between the mCTA predicted infarct and follow up infarct was 21.7 mL (limit of agreement (LoA): –41.0 to 84.3mL) in the 100 patients who had acute reperfusion (mTICI 2b/2c/3), and 3.4mL (LoA: –66 to 72.9mL) in the 44 patients who did not achieve reperfusion (mTICI 0/1). Amongst reperfused subjects, CCC was 0.4 [95%CI: 0.15–0.55, P<.01] and ICC 0.42 [95% CI: 0.18–0.50, P<.01]; in non-reperfused subjects CCC was 0.52 [95%CI: 0.2–0.6, P<.001] and ICC 0.6 [95% CI: 0.37–0.76, P<.001]. No difference was observed between the mCTA and CTP predicted infarct volume for the overall test cohort (P =.67).

**Conclusion:** Multiphase CT Angiography is able to predict infarct, penumbra and tissue perfusion, comparable to CT perfusion imaging.

## Introduction

Ischemic infarct core estimated using CT perfusion (CTP) at admission may be used in treatment decision making for patients with acute ischemic stroke (AIS).^1–4^ Classification of infarct core and penumbra is achieved using tissue perfusion estimates derived using a deconvolution algorithm from repeated serial imaging. The mismatch ratio between salvageable tissue (penumbra) volume and infarct core volume can be used for selecting patients presenting beyond 6 hours and up to 24 hours from last known well.^3^ CTP is limited by varying standardization of CTP parameter thresholds across different vendors, longer acquisition times and consequent susceptibility to patient motion, increased radiation dose, limited coverage (with some scanners) and the need for additional technical expertise to acquire the images.^5–7^

Multiphase computed tomographic angiography (mCTA) has been similarly used to select patients with AIS for endovascular therapy (EVT) in recent clinical trials.^8, 9^ Advantages of this technique compared to CTP are simpler image acquisition, lower radiation exposure, no additional contrast compared to single-phase CTA, and whole-brain time-resolved images of pial arteries and veins beyond an occlusion while also determining thrombus location, size, vessel patency and tortuosity.^10, 11^ Multiphase CTA imaging has not been as commonly used to predict ischemic tissue fate on a voxel by voxel basis, in the same way as CTP imaging. However, recent studies have demonstrated that mCTA can be used to predict tissue fate regionally, similar to CTP.^12–14^ An ability to harness the advantages of mCTA while producing brain maps that estimate tissue perfusion and predict tissue fate is likely to be of significant clinical utility.

We aimed to develop a machine learning based technique to estimate infarct core, penumbra and tissue perfusion in patients with acute ischemic stroke.

## Materials and Methods

Data were from the Prove-IT study (Precise and Rapid assessment of collaterals using multi-phase CTA in the triage of patients with acute ischemic stroke for IA Therapy), a multicenter study that acquired acute multimodal CT imaging including NCCT, multiphase CTA imaging (three phases), and CTP at baseline among ischemic stroke patients.^10,12^ This study was approved by the local institutional review board.

## Study Participants

Subjects who had (1) baseline non-contrast-enhanced CT (NCCT) and mCTA; (2) baseline CTP imaging with > = 8 cm z-axis coverage; (3) had reperfusion assessed on conventional angiography after thrombolysis treatment (intravenous tPA, endovascular therapy, or both) with the modified thrombolysis in cerebral infarction [mTICI]); and (4) had 24/36-hour follow-up imaging on diffusion MRI or NCCT were included in this analysis. Patient inclusion and exclusion are shown in Figure 1. We included 284 patients, of whom, 196 patients had acute reperfusion (mTICI 2b/2c/3) and 88 patients did not (mTICI 0/1).

**Figure 1.**
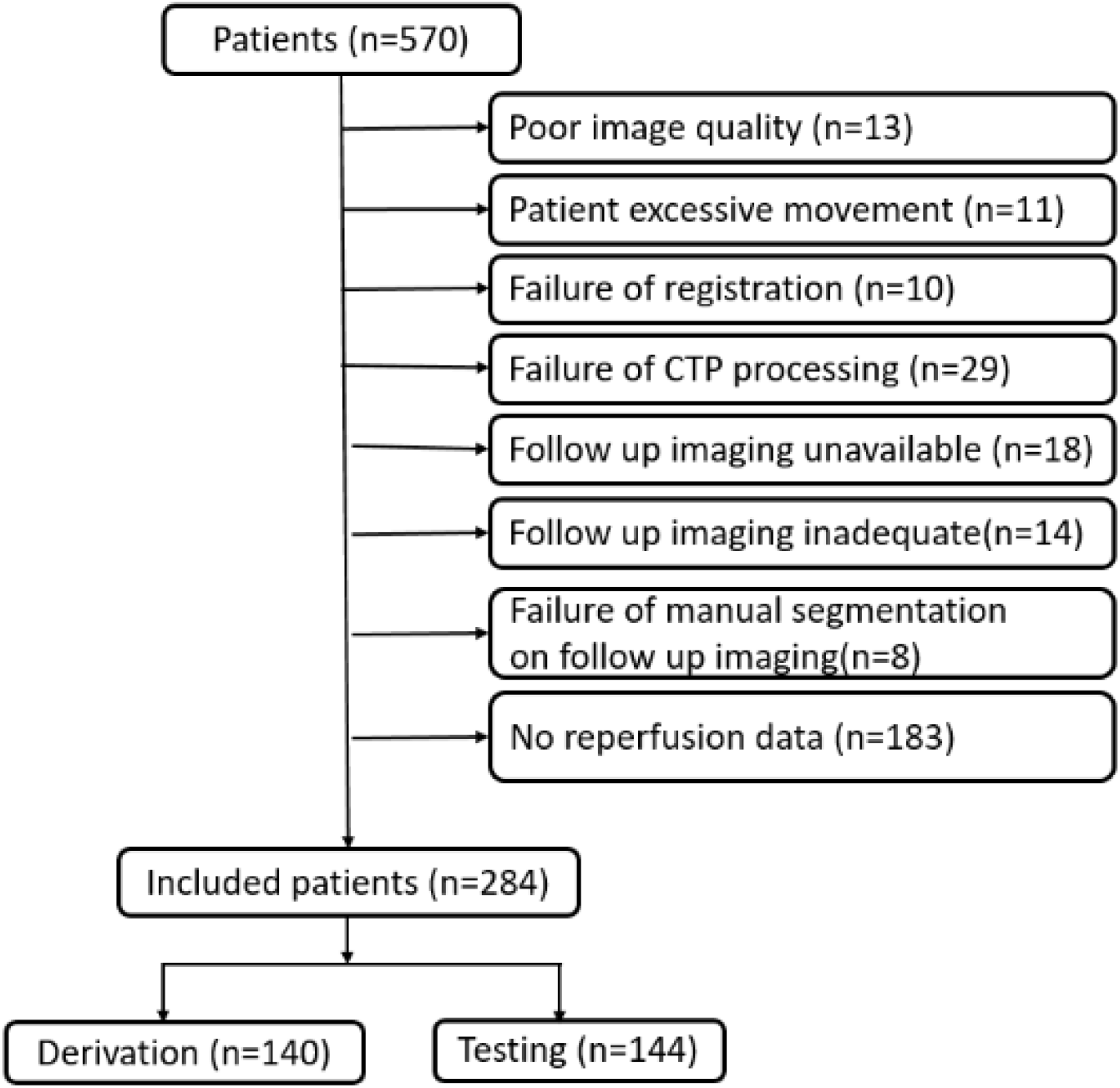
Patient inclusion chart.

## Image preprocessing

Each CTP study was processed using commercially available delay-insensitive deconvolution software (CT Perfusion 4D, GE Healthcare, Waukesha, WI). Absolute maps of cerebral blood flow (CBF, mL · min^−1^ · (100g)^−1^], cerebral blood volume (CBV, mL · (100 g)^−1^], and Tmax (seconds) were generated. Average maps were created by averaging the dynamic CTP source images. Time-dependent Tmax thresholds confirmed previously, were used to generate baseline CTP thresholded maps (perfusion volume).^6,7^

NCCT and mCTA images were first skull stripped.^15^ Three-phase CTA images were then aligned using rigid-body registration to account for patient movement. The aligned 3-phase CTA images were registered onto NCCT images using affine registration. Two radiologists (>5 years’ experience) used ITK-SNAP and consensus to manually delineate the infarct region on follow-up DWI/NCCT imaging.^16^ The follow-up images along with manual infarct segmentations and CTP average maps were registered onto NCCT images, thus bringing all images into the same image space. When registration was sub-optimal, manual refinement of the registered infarct segmentations was attempted. The NiftyReg tool was used for all image registration tasks.^17^

## Machine learning model

For this analysis, we defined *infarct core* as tissue that is infarcted on follow-up imaging even with reperfusion. *Penumbra* was defined as ischemic tissue that is not infarct core but infarcts on follow-up imaging when reperfusion is not achieved. These definitions of infarct core and penumbra are operational in context and not biological. The perfusion map used in this analysis was a Tmax map thresholded using previously published time dependent thresholds.^6, 7^

We developed three machine learning models: (1) Infarct model; (2) Penumbra model; (3) Perfusion model. A 2-stage training mechanism was developed to train two machine learning models to predict *infarct core* and *penumbra* respectively. The detailed training and testing strategy is shown in Figure 2. Of 88 patients without acute reperfusion (mTICI 0/1), 44 patients (35 for training and 9 for validation) were randomly selected to derive a random forest classifier at the first stage for prediction of follow-up infarction in the non-reperfused patients (Penumbra model), while the remaining 44 patients with mTICI 0/1 independent of the derivation cohort were used to test this derived Penumbra Model. Of those 196 patients with mTICI 2b/2c/3, 96 patient images (70 for training and 26 for validation) randomly selected were first processed by the 1^st^ stage Penumbra model, generating penumbra probability maps. These probability maps along with mCTA images were then used as inputs to derive the second random forest classifier at the second stage for infarct prediction (Infarct model) using follow up infarct manually segmented as reference standard, while the remaining 100 patients with mTICI 2b/2c/3 reperfusion independent of the derivation cohort were used to test the derived Infarct Model. The final predictions are shown as *infarct core* and *penumbra* where penumbra is defined as affected tissue from the penumbra model *minus* affected tissue from the infarct core model (Figure 3).

**Figure 2.**
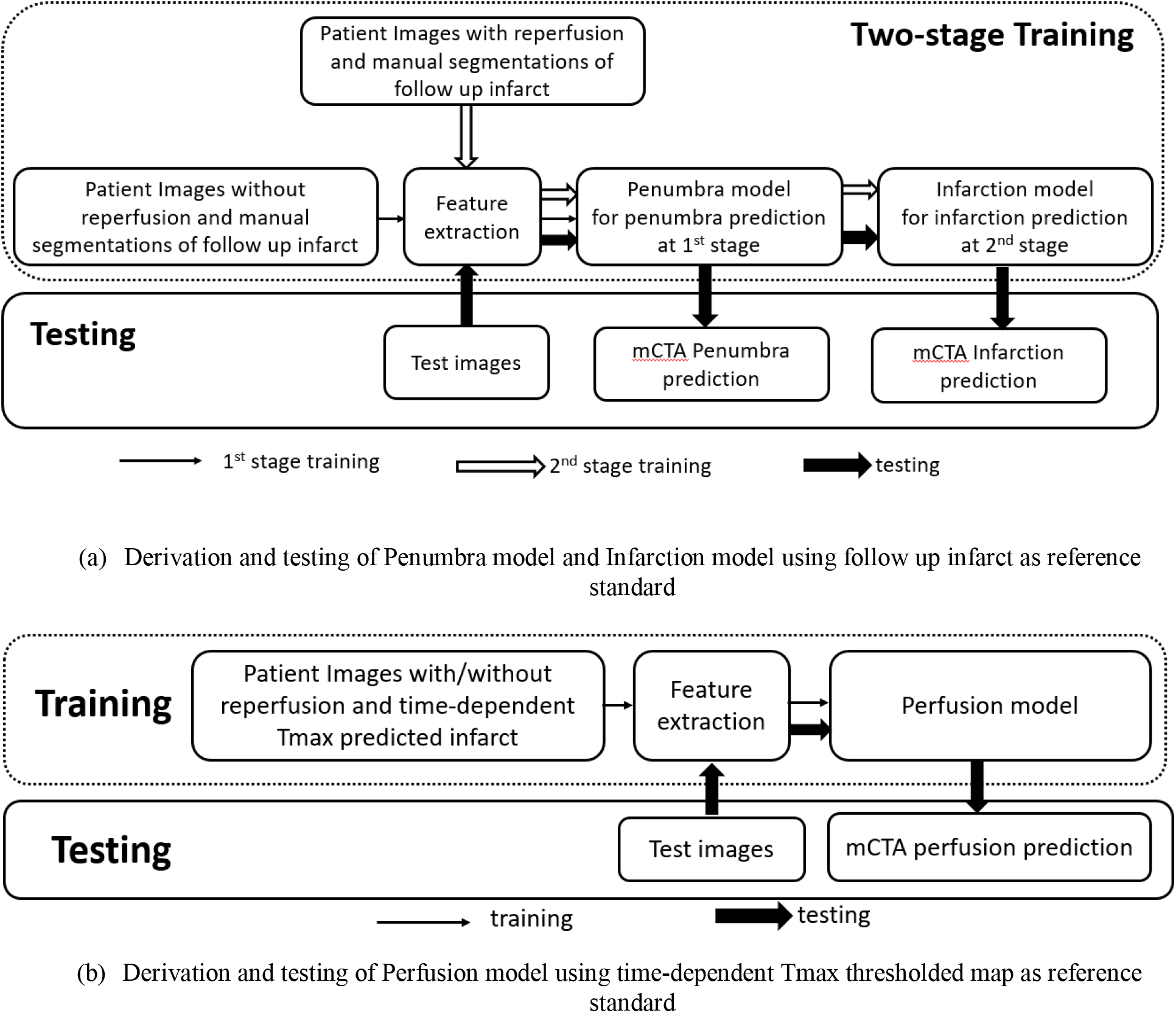
Training and Testing strategy of machine learning models to predict ischemic infarct, penumbra and perfusion status.

**Figure 3.**
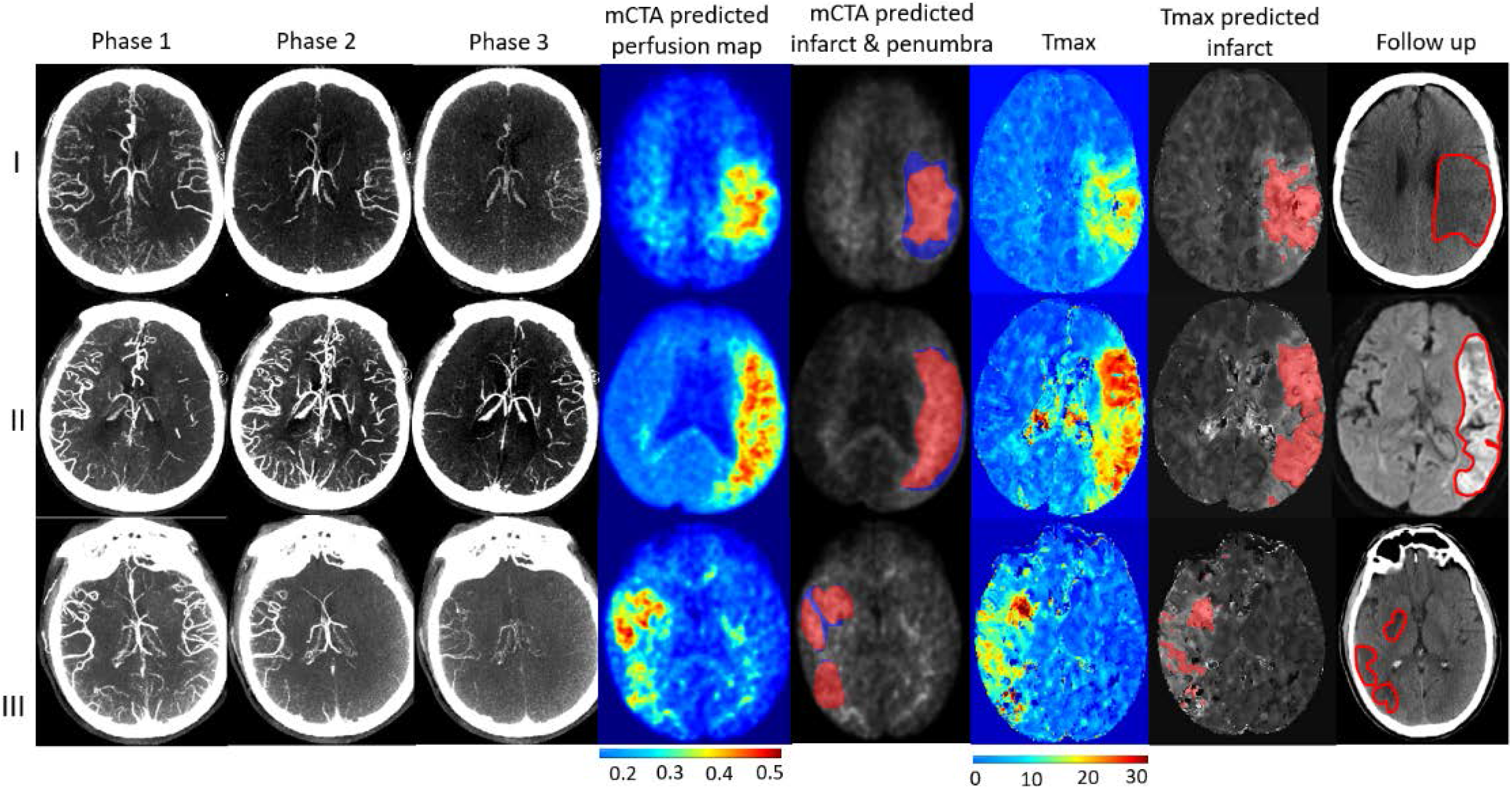
Multiphase CTA predicted infarct probability map compared to CTP (Tmax) map and follow up infarct. There are 3 examples, each labelled as per rows. Row I: patient with mTICI 2b, row II: patient with mTICI 1, and row III: patient with mTICI 3. Columns: mCTA phase 1 to 3, mCTA prediction perfusion maps, mCTA predicted infarct (red in column 5) and penumbra (blue in column 5) overlaid on the mCTA predicted perfusion map, CTP Tmax maps, time-dependent Tmax thresholds predicted infarct, infarct contoured in follow up imaging, respectively. The *penumbra* is shown as affected tissue from the Penumbra model *minus* affected tissue from the Infarct model.

In order to show the ability of mCTA to estimate tissue perfusion at baseline compared to CTP imaging, the 140 patient images used for training and validating the Penumbra and Infarct models were reused to train and validate the third random forest classifier (Perfusion model). For deriving and testing this model, time dependent Tmax thresholded maps were used as reference standard.^6,7^ The 144 images used for testing Penumbra and Infarct models independent on the derivation cohort were used to test the Perfusion model.

All three random forest models shared the same self-designed features as inputs. NCCT HU values were first subtracted from 3-phase CTA images, leading to a 3-point time intensity curve (TIC) for each voxel. Several features were extracted from the TIC for each voxel and used for deriving and testing the three random forest classifiers. The random forest classifiers derived from the training and validation dataset was then applied to the test cohort to generate a probability map for each patient. The probability map was then thresholded to generate the mCTA predicted volume.

## Statistical Methods

Expert contoured follow up lesion volume (Follow up infarct volume) were used as standard reference to evaluate mCTA predicted *infarct core* and *penumbra* volume for the test cohort. Time-dependent Tmax thresholded volumes (CTP volume) were used as standard reference to evaluate the mCTA perfusion volume for the test cohort. Bland-Altman plots were used to illustrate mean differences and limit of agreement (LoA) between mCTA predicted and follow up infarct volume, and CTP volume. Literal and relative volume agreement between mCTA predicted and follow up infarct volume, and CTP volume were also assessed using concordance correlation coefficient (CCC) and intra-class correlation coefficient (ICC), respectively. Spatial agreement between mCTA predicted volume and follow up infarct volume, and CTP volume was assessed using Dice similarity coefficient (DSC). Rank sum test was used to assess the difference between any non-normally distributed data. All statistical analyses were performed using MedCalc 17.8 (MedCalc Software, Mariakerke, Belgium) and Matlab (The MathWorks, Inc., United States). A two-sided alpha <0.05 was considered as statistically significant.

## Results

### Study Participants

Patient characteristics are summarized in Table 1. No differences were observed between the derivation and test cohorts (all P>.05).

**Table 1.** Patient characteristics.

| Characteristics | Derivation cohort<br>(N=140) | Test cohort<br>(N=144) | P value |
| --- | --- | --- | --- |
| Median age, year (IQR) | 73 (62–79) | 72 (62–80) | .73 |
| Sex, n(%) male | 80 (57) | 77 (53) | .56 |
| Median baseline NIHSS (IQR) | 17 (7–23) | 14 (6–18) | .12 |
| Median baseline ASPECTS (IQR) | 9 (8–10) | 9 (8–10) | .15 |
| Median onset-to-imaging time,<br>min (IQR) | 131 (94–226) | 139 (88–294) | .35 |
| Median imaging-to-reperfusion<br>time, min (IQR) | 90 (68–115) | 87 (64–125) | .97 |
| Median onset-to-reperfusion time,<br>min (IQR) | 245 (172–330) | 240 (181–377) | .71 |
| Median follow-up infarct volume,<br>mL (IQR) | 22.2(10.3-59.4) | 25.9 (10.1-60.6) | .60 |
| Site of occlusion, n(%) |  |  |  |
| ICA | 22(16) | 26(18) | .76 |
| MCA:M1 | 73(52) | 70(48) | .64 |
| other | 45(32) | 48(33) | .63 |

IQR, interquartile range; NIHSS, National Institutes of Health Stroke Scale. \*p<0.05.

### Accuracy of mCTA in predicting Follow up Infarct

Figure 3 demonstrates three examples of the mCTA prediction maps compared to CTP Tmax maps and follow up infarct. Figure 4(a) illustrates a Bland–Altman agreement plot between mCTA predicted *infarct core + penumbra* volume and follow up infarct volume for the 44 patients who did not receive acute reperfusion (mTICI 0/1) in the test cohort. The mean difference between this mCTA predicted *infarct core + penumbra* volume (median, 33.2; IQR, 20.6–53.2 mL) and follow up infarct volume (median, 26.8; IQR, 12.3–54.8 mL) was 3.4 mL (LoA: –66-72.9mL, P =.69). The CCC between the two volumes was 0.52 [95%CI: 0.2–0.6, P<.001] while the ICC was 0.6 [95% CI: 0.37–0.76, P<.001]. The median DSC between the mCTA predicted lesion and follow up infarct was 26.5% (IQR, 12.9–39.3%).

**Figure 4.**
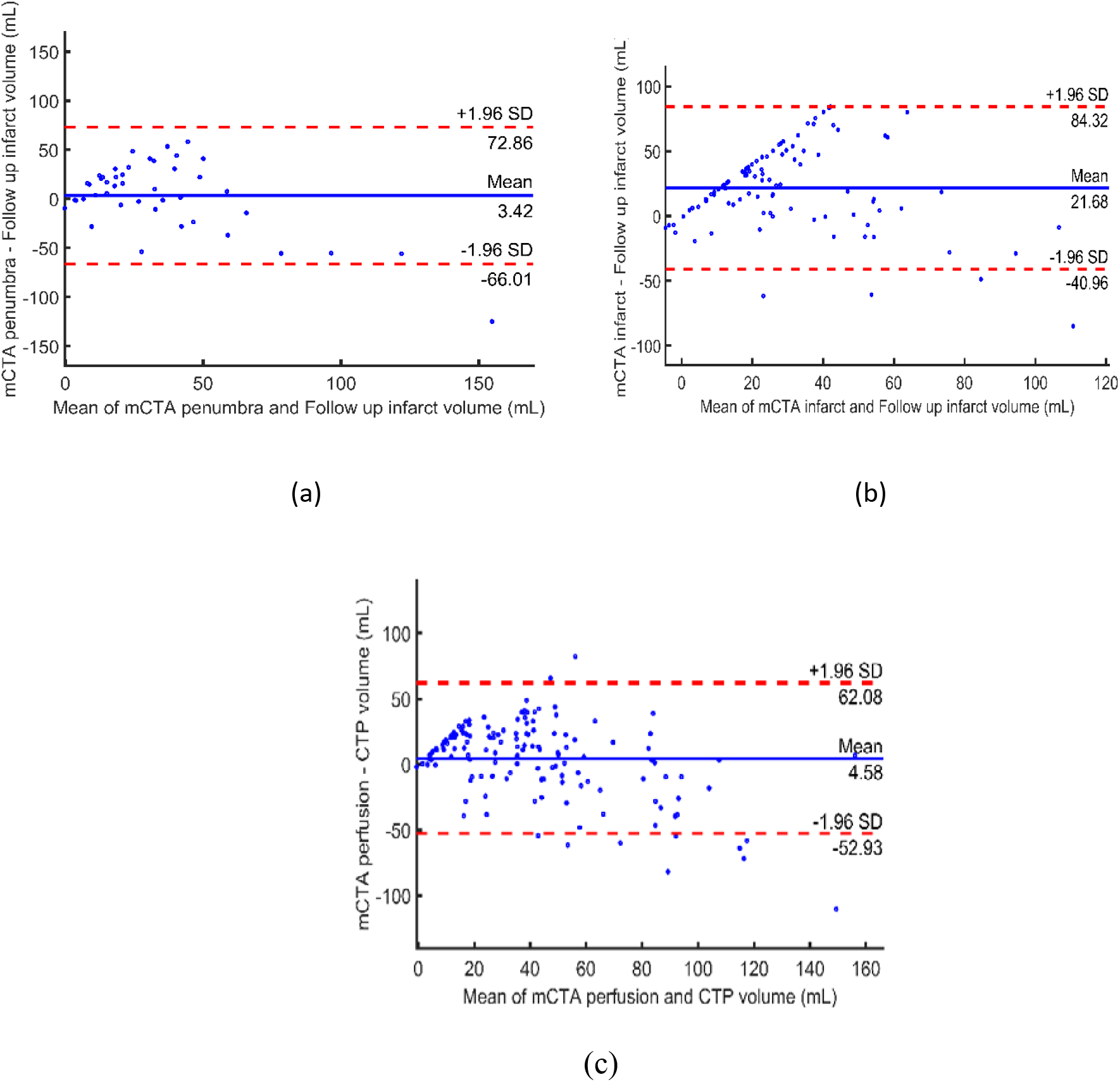
Bland-Altman plots of (a) mCTA penumbra volume predicted using Penumbra model vs. follow up infarct volume for 44 patients who did not achieve acute reperfusion (mTICI 0/1); (b) mCTA infarct volume predicted using Infarction model vs. follow up infarct volume for 100 patients who had acute reperfusion (mTICI 2b/2c/3); and (c) mCTA perfusion volume predicted using Perfusion model vs. time-dependent Tmax predicted infarct volume for all 144 patients in the test cohort.

Figure 4(b) illustrates a Bland–Altman agreement between the mCTA predicted infarct volume and follow up volume for 100 patients who achieved acute reperfusion (mTICI 2b/2c/3) in the test cohort. The mean difference between the mCTA infarct volume (median, 37; IQR, 23–58mL) and follow up volume (median, 26; IQR, 13–54mL) was 21.7 mL (LoA: –41.0-84.3mL, P =.48), CCC was 0.4 [95%CI: 0.15–0.55, P<.01] and ICC 0.42 [95% CI: 0.18–0.50, P<.01]. The median DSC between the mCTA infarct and follow up infarct was 24.7% (IQR, 13.8–30.4%).

The association between infarct volume predicted by the mCTA Infarct and Penumbra models and follow up infarct volume in the whole test cohort is shown in Table 2.

**Table 2.** Comparisons between infarct volumes predicted by the derived mCTA models and CTP vs. follow up infarct volume (median, 24.8; IQR, 10.5–58.8 mL) in the test cohort (n=144).

|  | mCTA *Infarct* and<br>*Penumbra model* | mCTA<br>*Perfusion model* | Time dependent<br>Tmax thresholded<br>model (CTP) | P value |
| --- | --- | --- | --- | --- |
| Predicted volume<br>(median [IQR], mL) | 37.3[21.3,57.8] | 40.5 [22.9,63] | 38.3 [15.0, 65.5] | .67 |
| Volume difference#<br>(mean [LoA], mL) | 21.7[-44, 86.3] | 20.4 [-51.3,92.1] | 22.3 [-42.6, 87.2] | .45 |
| DSC (median<br>[IQR], %) | 22.5[13.8,30.4] | 21.7[10.9,31.2] | 23.2 [13.9,33] | .55 |
| CCC [95% CI] | 0.43[0.18-0.58] | 0.41 [0.16-0.62] | 0.45 [0.32-0.54] | N/A |
| ICC [CI] | 0.5 [0.29-0.64] | 0.47 [0.3-0.56] | 0.54 [0.3-0.64] | N/A |

IQR, interquartile range; LoA, limit of agreement; CI, confidence interval; DSC: Dice similarity coefficient between the predicted volume and follow up infarct volume; CCC: concordance correlation coefficient; ICC: intra-class correlation coefficient. N/A: Not applicable.

### Accuracy of mCTA Predicting Perfusion Status

Figure 4(c) illustrates a Bland–Altman agreement between the mCTA predicted perfusion volume and CTP volume for 144 patients in the entire test cohort. The mean difference between the mCTA perfusion (median, 40.5; IQR, 22.9–59 mL) and CTP volume (median, 26.9; IQR, 6.7–56.7mL) was 4.6 mL (LoA: –53-62.1mL, P =.56), CCC was 0.63 [95%CI: 0.53–0.71, P<.01] and ICC was 0.68 [95% CI: 0.58–0.78, P<.001]. The median DSC between mCTA predicted perfusion and CTP volume was 40.5% (IQR, 25.7–52.7%).

The association between the volume predicted by mCTA perfusion model and follow up infarct volume, and between the time dependent Tmax thresholded predicted infarct volume and follow up infarct volume in the whole test cohort is shown in Table 2.

### Discussion

Multiphase CT angiography (mCTA) is a quick and easy-to-use imaging tool in selecting patients with acute ischemic stroke (AIS) for revascularization therapy.^10^ The developed machine learning technique described in this study shows that tissue status can be automatically predicted from the mCTA just as it is currently done using CT perfusion imaging. These results demonstrate that mCTA has similar ability to CTP imaging in predicting tissue fate. The developed ML technique could potentially help physicians make clinical decisions regarding acute stroke treatment, especially in hospitals without CTP capabilities.

When comparing mCTA predicted infarct volume with follow up infarct volume in patients who achieved acute reperfusion (mTICI 2b/2c/3), the mean volume difference of 21.7 mL, CCC of 0.4, and ICC of 0.42 are fair. The mCTA predicted infarct volume agrees better with follow up infarct volume in patients who did not achieve acute reperfusion (mTICI 0/1/2) with a mean volume difference of 3.4 mL, CCC of 0.52, and ICC of 0.6. DSCs between mCTA predicted infarct and penumbra and follow up volume are relatively low (less than 30%). However, accurate spatial quantification of infarction in patients with AIS is complicated and likely influenced by many pathophysiological factors, such as collateral status, tissue tolerance to ischemia/hypoxia, cerebral autoregulation, leukoaraiosis, fluctuation in blood pressure, hyperglycemia and time to reperfusion.^6,7,18^ All these factors likely lead to discrepancy between infarct volume predicted at baseline and follow up imaging. Of note, a recent paper from the HERMES group that used validated CTP software (i.e. RAPID, iSchemaView, Menlo Park, CA) showed similar DSC (median, 0.24; IQR, 0.15–0.37) between CTP predicted infarct volume and follow up infarct volume.^19^ When comparing mCTA predicted perfusion maps with CTP time dependent Tmax thresholded maps, the results show stronger agreement between the two measurements with a mean volume difference of 4.6 mL, CCC of 0.63, and ICC of 0.68. The median DSC of 40.5% between the mCTA predicted perfusion and CTP volume was also reasonable, suggesting good spatial overlap.

Imaging paradigms currently used in selecting patients with AIS for treatment include non-contrast CT, single-phase CTA or CTP. CTP, however, requires additional radiation and contrast and specific acquisition protocols that are different from NCCT and CTA. CTP is sensitive to patient motion, a feature that invalidates that tool in almost 10 to 25% of patients.^20^ Eleven patients were excluded from this study as CTP maps generated by the software were corrupted due to the excessive patient motion during acquisition (Figure 1). Multiphase CTA is potentially less prone to patient motion while being capable of predicting perfusion maps appropriately (Supplements: Figure 1). CTP also adds additional costs to the health system that include cost of the scan and resources allocated to training personnel to acquire the scan.^21, 22^ Multiphase CTA has minimal additional radiation, no additional contrast material, whole-brain coverage, relative insensitivity to patient motion and minimal additional cost to acquire over a single phase CTA.^10, 12,13^ The technique described here automates mCTA interpretation and provides physicians with tissue prediction as well as perfusion maps, just as CTP imaging does, thus potentially increasing their confidence in decision making (Table 2). Figure 3 shows an example of *penumbra* and *infarct core* (and mismatch) predicted using mCTA. Of note, physiological definitions of infarct and penumbra are different from the operational definitions used in this study. Unlike conventional mCTA but similar to CTP, the technique described in this study is capable of detect smaller perfusion lesions in the entire brain including the posterior circulation (Supplements: Figure 2).

A strength of the developed machine learning technique is that it does not rely on deconvolution algorithms, which plays an essential role in current CTP processing. Although deconvolution methods can appropriately model perfusion status, the introduction of physiological variations in arterial delivery of contrast, the effects of collateral flow, and venous outflow components of cerebral perfusion, greatly increase the computational complexity.^23, 24^ The number of variables and the algorithms used to calculate these variables results in variability in generating CTP threshold values for estimating infarct and penumbra across different vendor software. Additionally, numerical solutions to deconvolution greatly relies on accurate selection of artery input function (AIF), a parameter that is case dependent and sensitive to noise, especially given the low signal to noise ratio of perfusion images, even when preprocessing, such as motion correction, temporal and spatial smoothing, and deconvolution regularization are applied.^25,26^ The deconvolution free approach developed in this study can be easily integrated into any imaging paradigm using NCCT and mCTA as a post-processing step, potentially obviating the need for CTP.

This study has several limitations. First, of 284 patients used for this study, 166 patients had follow up NCCT imaging. The variability of manual infarct segmentations on NCCT may increase the complexity of training machine learning models and decrease the validity of these results. Second, our current models were derived using imaging information only. Previous studies have suggested that adding clinical information, such as time^8, 9^ and patient characteristics^28^ to the models may improve prediction accuracy. Integrating this information into the models should be further investigated^29^. Third, the machine leaning models were derived from a limited number of training data. Increasing the amount and diversity of training data would help to improve the accuracy and robustness of the model, thereby accounting for variance in acquisition across patients and scanners. Fourth, validating the models on larger datasets is necessary in order to translate these into clinical practice.

In conclusion, infarct core, penumbra, and perfusion status can be automatically predicted from multiphase CTA imaging using machine learning. This technique shows comparable accuracy to CT perfusion imaging in measuring tissue status in patients with acute ischemic stroke. This work has the potential of assisting physicians in making treatment decisions in clinical settings.

## Data Availability

The data is not publicly available.

